# Predicting county-level diagnosed diabetes prevalence in the United States using explainable gradient boosting and geographic interpretation

**DOI:** 10.64898/2026.06.23.26356400

**Authors:** Yussif Yahaya, Sagor Khan, Priyanka Rani Saha, Md Al Amin Meia

## Abstract

Diagnosed diabetes affects approximately 38.4 million Americans, but its burden is not evenly distributed across U.S. counties. Existing machine-learning studies have mainly focused on individual risk prediction using biometric, clinical, or survey variables. These approaches are less suited to explaining why diagnosed diabetes prevalence differs geographically across counties. We developed an explainable gradient-boosting framework for predicting county-level diagnosed diabetes prevalence across 2,957 U.S. counties using an ecological cross-sectional design. The analysis integrated food-environment, socioeconomic, occupational, demographic, health-behavior, and clinical indicators from five public data sources. Four regression models were compared: Elastic Net, Random Forest, XGBoost, and LightGBM. LightGBM was selected as the primary model based on validation-set RMSE and interpreted using SHAP TreeExplainer. The validation-selected LightGBM model achieved a held-out test RMSE of 0.423 percentage points, *R*^2^ = 0.964, and MAPE = 2.76%. Although XGBoost achieved a lower test RMSE of 0.399 and *R*^2^ = 0.968, it was retained as a secondary benchmark because primary-model selection was based only on validation performance. A sensitivity model using only structural and contextual predictors, and excluding CDC PLACES health-behavior and clinical covariates, retained substantial predictive performance (*R*^2^ = 0.827). Poverty rate was the most frequent dominant positive structural SHAP contributor nationally (*n* = 772 counties, 26.1%), followed by food insecurity rate (*n* = 707, 23.9%), Supplemental Nutrition Assistance Program (SNAP) participation rate (*n* = 316, 10.7%), unemployment rate (*n* = 224, 7.6%), and median household income (*n* = 178, 6.0%). Residual Moran’s *I* decreased from 0.665 to 0.069 after model fitting. Explainable machine learning using public county-level data can characterize geographic variation in diagnosed diabetes prevalence. County-level SHAP maps may support local hypothesis generation, but should be interpreted as explanations of model predictions rather than causal effects.

**Author Summary:** Diabetes prevalence differs substantially across U.S. counties, but the local conditions associated with these differences are not always clear. We combined public county-level data on diagnosed diabetes, health behaviors, poverty, unemployment, food insecurity, food access, demographic composition, rurality, and occupational structure for 2,957 counties. We compared four statistical and machine-learning models. LightGBM was selected as the primary model because it had the lowest validation-set RMSE, while XGBoost was retained as a secondary benchmark because it achieved slightly lower error on the held-out test set.

We then used SHapley Additive exPlanations (SHAP) to examine how county characteristics contributed to the LightGBM predictions. Physical inactivity was the most influential predictor overall. Among structural and contextual predictors, poverty, food insecurity, Supplemental Nutrition Assistance Program (SNAP) participation, unemployment, and median household income frequently made the largest positive contributions to predicted diagnosed diabetes prevalence.

Our findings show how explainable machine learning can turn public health data into geographically specific model explanations. These maps may help identify counties and structural conditions needing further investigation, but they describe predictive patterns and should not be interpreted as causal effects or intervention recommendations.

## 1 Introduction

Diagnosed diabetes, most of which reflects type 2 diabetes mellitus (T2DM) among adults, remains a major chronic disease burden in the United States. The Centers for Disease Control and Prevention National Diabetes Statistics Report estimated that 38.4 million Americans, representing 11.6% of the population, were living with diabetes in 2021, including 8.7 million undiagnosed cases [1]. The prevalence of diabetes has nearly doubled over the past three decades, partly in relation to rising obesity, physical inactivity, and population aging [1]. The economic burden is also substantial: the American Diabetes Association estimated that diagnosed diabetes cost $412.9 billion in 2022, including $306.6 billion in direct medical costs and $106.3 billion in lost productivity, with medical expenditures among people with diagnosed diabetes estimated to be 2.6 times higher than among those without diabetes [2].

This burden is geographically uneven. County-level variation reflects differences in food access, economic conditions, rurality, healthcare infrastructure, and demographic context. Chen et al. [3] reported that the East South Central census division, including Alabama, Mississippi, Kentucky, and Tennessee, sustained some of the highest diagnosed diabetes prevalence rates during 2011–2021. Racial, ethnic, and rural–urban disparities also shape these patterns: diagnosed diabetes prevalence was highest among American Indian and Alaska Native adults, non-Hispanic Black adults, and adults of Hispanic origin compared with non-Hispanic White adults [1], and rural diabetes prevalence exceeded urban prevalence nationally and in many states after demographic adjustment [4]. Together, these patterns suggest that county-level structural and contextual conditions may be associated with geographic differences in diabetes burden beyond individual clinical risk factors.

Machine learning has been widely used for diabetes risk prediction, with tree-based ensemble models often performing well compared with traditional regression. Studies using the CDC Behavioral Risk Factor Surveillance System and the National Health and Nutrition Examination Survey have reported ROC-AUC values above 0.88, with XGBoost and Random Forest frequently among the better-performing algorithms [5, 6]. However, much of this work has focused on individual-level risk classification using biometric, clinical, or survey variables such as body mass index, glycated hemoglobin, blood pressure, age, and self-reported socioeconomic characteristics. These models are useful for identifying who may be at elevated risk, but they are less suited to explaining why diagnosed diabetes prevalence is concentrated in particular counties or which local structural conditions are most strongly associated with that concentration. BRFSS-based studies have also acknowledged that environmental and neighborhood-level conditions, including dietary quality and healthcare access, may be missing from standard prediction models [7].

A related geospatial epidemiology literature has examined how place-based conditions are associated with diabetes burden. Food access, fast-food density, grocery access, poverty, and neighborhood socioeconomic disadvantage have been linked to diabetes outcomes in spatial regression and geographic information system studies. For example, Oh et al. [8] applied spatial error models and geographically weighted regression to 3,108 contiguous U.S. counties and reported strong spatial clustering of diabetes prevalence (Global Moran’s *I* = 0.593, *p <* 0.0001), with food desert exposure positively associated with diabetes in southern counties. Bleich et al. [9] similarly found that food swamps remained associated with diabetes outcomes in North Carolina after accounting for spatial autocorrelation. These studies support the importance of local food environments, but many classical spatial models have limited ability to capture nonlinear relationships while also producing locally interpretable prediction outputs. Reviews also note that the relationships among food insecurity, neighborhood food environments, and health disparities remain incompletely understood [10, 11].

Explainable artificial intelligence offers one way to connect predictive modeling with public-health interpretation. SHapley Additive exPlanations (SHAP) have been used in diabetes research to identify influential predictors and clarify how model outputs differ across population groups. Kambara et al. [12], for example, applied SHAP to models trained on the NIH *All of Us* cohort and found that income, waist circumference, and education contributed to model explanations of higher T2DM prevalence among Black or African American participants. However, these explanations were based mainly on individual-level survey and clinical variables. Less attention has been given to SHAP-based interpretation of externally linked county-level structural measures, including food access, food retail environment, socioeconomic disadvantage, and local health-behavior patterns. County-level interpretability is important because public-health planning often requires evidence about where disease burden is concentrated and which local conditions contribute most strongly to predicted prevalence.

The present study addresses this gap by developing an explainable machine-learning framework for predicting county-level diagnosed diabetes prevalence in the United States. We linked county-level diagnosed diabetes prevalence with publicly available indicators of food access, food environment, socioeconomic conditions, demographic composition, rurality, and health behaviors using county Federal Information Processing Standards codes. Gradient-boosting models were used to capture nonlinear associations among predictors, and SHAP analysis was applied to identify both overall feature importance and county-specific contributors to predicted diagnosed diabetes prevalence. Specifically, this study aimed to:

1. compare machine-learning models for predicting county-level diagnosed diabetes prevalence among U.S. counties with available linked data;
2. identify the strongest structural, food-environment, socioeconomic, demographic, and health-behavior predictors of county-level diagnosed diabetes prevalence using SHAP-based feature importance; and
3. map SHAP-derived county-level dominant positive structural contributors to support local hypothesis generation and geographically informed diabetes-prevention planning.

## 2 Methods

### 2.1 Study Design and Unit of Analysis

This study used a county-level ecological cross-sectional design. The county was chosen as the unit of analysis because food access, socioeconomic disadvantage, rurality, and local health behaviors can vary substantially within a single state and are better measured at the sub-state level [13].

The primary outcome was 2023 county-level age-adjusted diagnosed diabetes prevalence from the 2025 CDC PLACES release, based on 2023 BRFSS data. Predictors came from the closest available pre-outcome or contemporaneous sources: the 2019 USDA Food Access Research Atlas, the 2019–2023 ACS 5-year estimates, and the 2019–2020 USDA Food Environment Atlas. Because this study was designed for cross-sectional geographic prediction rather than causal inference, predictor years were selected to maximize county coverage and temporal proximity to the 2023 outcome, while avoiding future information where possible. The temporal gap between the 2019 FARA release and the 2023 outcome reflects the slow-changing nature of food-environment and socioeconomic conditions, consistent with prior county-level ecological studies.

The analysis had six main steps: (i) county-level data assembly and harmonization; (ii) population-weighted aggregation of tract-level food-access indicators to counties; (iii) feature engineering and missing data treatment; (iv) model training and comparison; (v) SHAP-based geographic interpretation; and (vi) spatial autocorrelation and sensitivity analyses.

### 2.2 Data Sources and Variable Selection

Five public data sources were assembled and linked using the five-digit county Federal Information Processing Standards (FIPS) code as the merge key. Table 1 lists each source, the variables extracted, and the index year used.

**Table 1:**
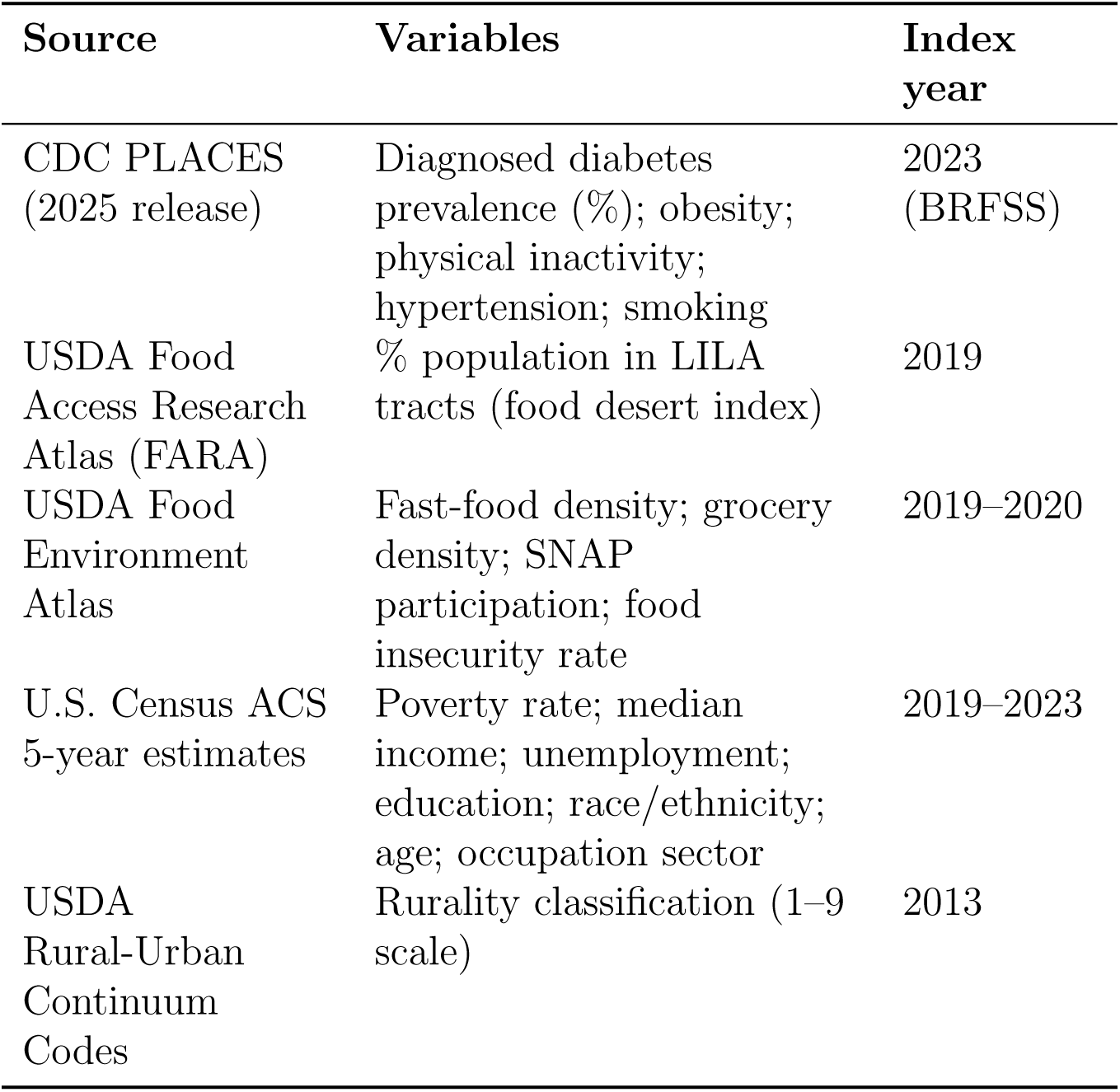
Data sources, variables, and index years.

**Table 2:** Performance of the four candidate models on the validation set used for model selection and on the held-out test set. Both sets contained 444 counties. Bootstrap 95% confidence intervals for test-set RMSE and MAE were estimated using 1,000 resamples.

| Model | Validation set | Held-out test set |  |  |  |
| --- | --- | --- | --- | --- | --- |
| | RMSE | RMSE (95% CI) | MAE (95% CI) | MAPE (%) | $R^2$ |
| LightGBM<sup>a</sup> | **0.442** | 0.423 (0.374–0.482) | 0.311 (0.287–0.338) | 2.76 | 0.964 |
| XGBoost | 0.446 | **0.399 (0.350–0.453)** | **0.290 (0.265–0.318)** | **2.56** | **0.968** |
| Random Forest | 0.562 | 0.538 (0.473–0.615) | 0.396 (0.362–0.430) | 3.53 | 0.941 |
| Elastic Net | 0.579 | 0.585 (0.538–0.637) | 0.454 (0.424–0.488) | 4.09 | 0.930 |

<sup>a</sup>Validation-selected primary model used for SHAP interpretation. Bold indicates the best numerical performance within each column. Pairwise Wilcoxon signed-rank tests compared paired absolute prediction errors on the held-out test set; all comparisons were statistically significant at $p < 0.01$ .

#### 2.2.1 Outcome variable

The outcome was county-level age-adjusted diagnosed diabetes prevalence per 100 adults, obtained from the 2025 CDC PLACES release. CDC PLACES produces small-area estimates by applying multilevel regression and post-stratification (MRP) to 2023 Behavioral Risk Factor Surveillance System (BRFSS) data [14]. Public surveillance data report diagnosed diabetes without separating diabetes subtypes. The outcome was therefore treated as a county-level proxy for type 2 diabetes burden, because type 2 diabetes accounts for approximately 90–95% of adult diabetes cases nationally [1].

#### 2.2.2 Health-behavior and clinical covariates

County-level obesity, physical inactivity, hypertension, and smoking prevalence were also taken from CDC PLACES. These variables are established correlates of diabetes burden and were included as covariates. Because they come from the same BRFSS/MRP process as the outcome, a structural-and-contextual-predictors-only sensitivity model was estimated to check whether model performance depended mainly on these health-behavior estimates rather than the structural, contextual, food-environment, and socioeconomic variables of primary interest (Sensitivity Model S1, Table 4).

#### 2.2.3 Food desert index

The USDA FARA defines low-income, low-access (LILA) census tracts as areas meeting both an income threshold and a supermarket-distance threshold. Low-income tracts have a poverty rate of at least 20% or a median family income at or below 80% of the statewide median. Low-access tracts are those where at least 500 people or 33% of the tract population live more than 0.5 miles from the nearest supermarket in urban areas, or more than 10 miles in rural areas [15]. Because FARA is published at the census-tract level, tract values were aggregated to counties using a population-weighted formula:

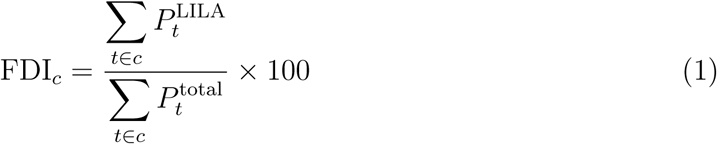

where FDI*_c_* is the county food desert index, *P*_t_^LILA^ is the number of residents in tract *t* meeting the LILA definition, *P*_t_^total^ is the tract population, and the sum covers all tracts within county *c*.

#### 2.2.4 Food Environment Composite Index

Three USDA Food Environment Atlas indicators were used to construct a composite food-environment measure: fast-food restaurant density per 1,000 population, grocery store density per 1,000 population, and SNAP participation rate. The three indicators were standardized to *z*-scores and combined using principal component analysis (PCA). The first principal component met Kaiser’s criterion (eigenvalue = 1.188 *>* 1.0) and explained 39.6% of total variance; it was therefore retained as the Food Environment Composite Index (FECI). Higher FECI scores reflected greater grocery store density and lower SNAP participation rates, representing a food retail access gradient.

The FECI was constructed before data partitioning using PCA fitted on the full analytic sample. This limited unsupervised preprocessing leakage is disclosed in the Limitations section. The FECI was retained in the primary model as a theoretically motivated a priori composite of food retail access. Because the FECI overlaps with its component indicators, we evaluated sensitivity specifications in which the FECI was excluded while the individual food-environment indicators were retained (Sensitivity Models S3 and S6, Table 4). If the first component had not met Kaiser’s criterion, the three indicators would have been entered separately in the primary model. This approach follows [13].

#### 2.2.5 Occupational sector and rurality

Occupational data came from ACS 5-year county estimates [16, 17]. We extracted the share of the civilian employed population in three NAICS macro-sectors: agriculture, forestry, fishing, and hunting (Sector 11); manufacturing (Sectors 31–33); and accommodation and food services (Sector 72). These sectors were selected because they may reflect local income instability, food access conditions, and occupational health exposures relevant to diabetes burden [18].

County rurality was measured using the 2013 USDA Rural–Urban Continuum Codes (RUCC), the release closest to the study period [19]. Because rurality classification changes slowly, RUCC was treated as a stable county contextual variable. A sensitivity model excluding RUCC was also estimated (Sensitivity Model S7, Table 4).

### 2.3 County-Level Data Harmonization

All datasets were linked using the five-digit county FIPS code. FIPS codes were stored as zero-padded character strings to preserve leading zeros and avoid errors from numeric conversion, which particularly affects states whose codes begin with zero. Tract-level FARA records were assigned to counties using the first five digits of the tract FIPS code, then aggregated using the population-weighted formula in Eq. 1.

One county was excluded due to missing outcome data, yielding a final analytic sample of 2,957 U.S. counties. The United States comprises 3,143 county and county-equivalent units; the difference between that total and our analytic sample reflects county-equivalent units and county records that were not consistently available or linkable across all five data sources via FIPS code. These include county-equivalent units such as independent cities in Virginia, Alaska borough subdivisions, and U.S. territories. Pennsylvania and Kentucky county records were not available in the CDC PLACES 2025 county-level file used for this analysis and were therefore excluded during outcome linkage. The analysis was restricted to counties and county-equivalent units with complete outcome linkage across CDC PLACES and the assembled predictor datasets. Counties shown in gray in the dominant-contributor map indicate county geometries present in the map shapefile but not included in the final analytic sample.

Missing predictor values were handled using iterative multivariate single imputation implemented with scikit-learn’s IterativeImputer with 10 iterations. The outcome variable was not included in the imputation model. A full summary of missing predictor values is provided in S1 Table. To avoid leakage from the validation and held-out test partitions, the imputer and min–max scaler were fitted using the training partition only and then applied without modification to the validation and held-out test partitions. During cross-validation within the training partition, these supervised preprocessing steps were performed within the corresponding training folds.

The Food Environment Composite Index was constructed before data partitioning using principal component analysis fitted on the full analytic sample. This limited unsupervised preprocessing leakage is disclosed in the Limitations section and was evaluated through sensitivity analyses excluding the FECI.

### 2.4 Machine-Learning Model Development

Four regression models were trained to predict county-level diagnosed diabetes prevalence: Elastic Net [20], Random Forest [21], XGBoost [22], and LightGBM [23]. Elastic Net was included as a regularized linear benchmark, Random Forest as a nonlinear bagging benchmark, XGBoost as a gradient-boosting comparator, and LightGBM as a candidate gradient-boosting model for primary interpretation. All four models used the same feature set and a fixed random seed (seed = 42).

Hyperparameters were tuned using cross-validated grid search within the training partition. Elastic Net tuning considered the regularization strength and *ℓ*_1_ mixing ratio; Random Forest tuning considered the number of trees, maximum depth, minimum samples per leaf, and maximum features; XGBoost and LightGBM tuning considered tree depth, learning rate, number of estimators, subsampling, column sampling, and tree-complexity parameters. The full hyperparameter grids and selected values for all four models are reported in S3 Table.

The primary prediction model was selected using RMSE on the validation partition only. After primary-model selection, the selected model was evaluated once on the held-out test set. XGBoost was retained as a secondary comparator to assess the robustness of predictive performance and SHAP-based geographic interpretations across gradient-boosting implementations. Both XGBoost and LightGBM are compatible with SHAP TreeExplainer, which estimates feature contributions efficiently for tree ensembles [24].

### 2.5 Model Validation and Performance Metrics

Counties were split into training (70%), validation (15%), and held-out test (15%) partitions stratified by Census region to preserve representation from the Northeast, Midwest, South, and West in each partition. No held-out test observations were used during preprocessing, hyperparameter tuning, or primary-model selection. The held-out test set was evaluated only after the primary model had been selected using validation-set RMSE.

Performance was measured using root mean squared error (RMSE), mean absolute error (MAE), mean absolute percentage error (MAPE), and *R*^2^. Bootstrap resampling with 1,000 iterations was used to estimate 95% confidence intervals for RMSE and MAE on the held-out test set. As a secondary non-parametric comparison, paired absolute prediction errors from competing models were compared using the Wilcoxon signed-rank test.

The primary model was selected exclusively by validation-set RMSE. The validation-selected model was then evaluated once on the held-out test set. The same selected primary model was used for SHAP interpretation, whereas XGBoost was retained as a secondary gradient-boosting comparator for cross-model robustness analyses.

Geographic generalizability was assessed through region-held-out validation. In this analysis, each Census region was held out in turn while the model was trained on counties from the other three regions. Within each regional fold, imputation and scaling were refitted using only the training regions and then applied to the held-out region.

### 2.6 Sensitivity Analyses

Nine sensitivity analyses were conducted to assess whether the main findings were robust to alternative predictor sets, variable constructions, preprocessing procedures, and modeling choices. All sensitivity specifications used the validation-selected LightGBM architecture and the same selected hyperparameters as the primary model. Models were evaluated on the same held-out test set and were treated as robustness checks rather than alternative candidates for primary-model selection.

The sensitivity analyses included nine robustness checks. S1 used only structural and contextual predictors and excluded CDC PLACES health-behavior and clinical covariates. S2 used only the health-behavior and clinical covariates. S3 and S6 excluded the FECI and differed only in whether preprocessing was refitted. S4 used the collinearity-reduced predictor set from the iterative VIF assessment. S5 replaced the continuous food desert index with a binary high-food-desert indicator. S7 excluded rurality, S8 added Census-region indicators, and S9 used the 10 highest-ranked predictors based on training-partition SHAP values. Full specifications are provided in S4 Table.

### 2.7 SHAP Explainability and Geographic Interpretation

Model interpretation used TreeExplainer from the shap library [24], applied to the validation-selected LightGBM model. SHAP values quantify how each predictor contributes to an individual model prediction relative to the model baseline. In this study, they were interpreted as explanations of model predictions, not as estimates of causal effects.

After model selection, SHAP explanations were generated from the LightGBM model fitted on the training partition (*n* = 2,069 counties). Training-fitted imputation and min–max scaling transformations were applied without modification to the validation and test partitions, and the fitted model was then applied to all 2,957 analytic counties. SHAP values for training counties were therefore in-sample explanations, whereas values for validation and test counties were out-of-sample explanations. The model was not refitted on the full analytic sample before SHAP computation.

We produced four SHAP summaries. Global mean absolute SHAP values were used to rank predictor importance. County-level SHAP maps showed where selected structural predictors contributed positively or negatively to model predictions. Regional signed SHAP summaries described broad directional patterns across Census regions. Finally, for each county, we identified the structural predictor with the largest positive SHAP value. This dominant positive structural contributor analysis describes the fitted model prediction and should not be interpreted as a causal determinant or as the preferred intervention target.

To assess consistency across gradient-boosting implementations, we compared SHAP explanations from the primary LightGBM model with those from the secondary XGBoost model. Agreement was evaluated using Spearman’s rank correlation between global mean absolute SHAP rankings, overlap among the five highest-ranked predictors, the percentage of counties assigned the same dominant positive structural contributor, and Pearson’s correlation between county-level predictions.

Collinearity was assessed using variance inflation factors calculated from the imputed training partition and reported in S2 Table. Because tree-based models do not estimate regression coefficients, VIF screening was not used to remove predictors from the primary LightGBM model. However, correlated predictors can redistribute SHAP attribution across related variables. Therefore, a collinearity-reduced sensitivity model was evaluated as S4. Full details of the VIF assessment and the S4 predictor-removal sequence are provided in the supporting information.

### 2.8 Spatial Autocorrelation Diagnostics

Spatial autocorrelation was assessed for both the observed outcome and the residuals from the validation-selected LightGBM model. Global Moran’s *I* was calculated for county-level diagnosed diabetes prevalence to quantify overall spatial clustering [25]. Global Moran’s *I* was then calculated for LightGBM residuals, defined as observed minus predicted prevalence, to assess whether geographically structured variation remained after model fitting. Statistically significant residual autocorrelation was interpreted as evidence that some spatially patterned variation remained unexplained by the included predictors.

The validation-selected LightGBM model fitted on the training partition was applied to all 2,957 analytic counties before residual spatial diagnostics were conducted. Residuals for training counties were therefore in-sample, whereas residuals for validation and test counties were out-of-sample. The residual spatial analysis was treated as a descriptive post-modeling diagnostic rather than as an independent test of predictive performance.

Spatial analyses included 2,912 of the 2,957 analytic counties; 45 counties were excluded because their county identifiers could not be matched to the geometries used to construct the spatial weights. Queen contiguity was used as the primary spatial-weight definition, with rook contiguity and five-nearest-neighbor weights (*k* = 5) evaluated as sensitivity specifications. All spatial weights were row-standardized, and statistical significance for Global Moran’s *I* was assessed using 999 permutations.

Local Indicators of Spatial Association were calculated using row-standardized queen-contiguity weights to identify high–high and low–low clusters of observed diagnosed diabetes prevalence, as well as high–low and low–high spatial outliers [26]. Local statistical significance was evaluated using 999 permutations and an unadjusted threshold of *p <* 0.05. Because thousands of local tests were conducted simultaneously, the LISA results were interpreted as exploratory. Spatial lag and spatial error regression models were not used as the primary modeling approach because the study objective was county-level prediction with SHAP-based geographic interpretation rather than estimation of spatial regression coefficients.

### 2.9 Computational Environment

Data assembly, harmonization, feature engineering, model training, SHAP analysis, spatial diagnostics, sensitivity analyses, and figure generation were performed in Python using reproducible analysis scripts. Initial preprocessing was conducted locally, and model training and post-modeling analyses were conducted on the University of Texas Rio Grande Valley CRADLE high-performance computing cluster. Key packages included scikit-learn, XGBoost, LightGBM, SHAP, GeoPandas, libpysal, and esda. Complete package versions, environment specifications, processed datasets, model outputs, and analysis scripts are provided with the archived code and data described in the Data Availability statement.

## 3 Results

### 3.1 Sample Description and Spatial Distribution

Of 3,143 U.S. counties and county-equivalent units, 2,958 records were initially retained after linking the CDC PLACES outcome file with the assembled predictor datasets. One record lacked a valid diagnosed diabetes prevalence estimate and was excluded, yielding a final analytic sample of 2,957 counties. Pennsylvania and Kentucky had no county-level records in the CDC PLACES file used for this analysis and were therefore absent from the analytic sample. Counties from all four Census regions were represented: South (1,301 counties, 44.0%), Midwest (1,055, 35.7%), West (450, 15.2%), and Northeast (151, 5.1%).

County-level diagnosed diabetes prevalence ranged from 6.0% to 23.8%, with a mean of 11.2% (SD = 2.6%). The food desert index ranged from 0% to 100%, with a mean of 25.5% (SD = 25.1%) and a median of 21.4%. Using the study-defined threshold of a food desert index *≥* 20%, 1,644 counties (55.6%) were classified as high food-desert counties. The Food Environment Composite Index had a mean of 0.00 (SD = 1.09), with higher values indicating greater grocery store density and lower SNAP participation rates. Grocery store density had the highest predictor missingness at 29.6%. Missing grocery-density values were treated as unavailable observations rather than as true zero values. Missing predictor values were handled using the prespecified iterative multivariate single imputation procedure. All other predictors had missingness below 16% and were processed using the same procedure. A complete summary of predictor missingness is provided in S1 Table.

Spatial analyses included 2,912 of the 2,957 analytic counties. Forty-five counties were excluded because their county identifiers could not be matched to the geometries used to construct the spatial weights. Global Moran’s *I* for county-level diagnosed diabetes prevalence was *I* = 0.665 (permutation *p* = 0.001, queen contiguity), indicating strong positive spatial clustering. This pattern was robust to alternative spatial-weight definitions, including rook contiguity (*I* = 0.667) and five-nearest-neighbor weights (*I* = 0.677), both with permutation *p* = 0.001. Local Indicators of Spatial Association identified high–high clusters primarily in the South, particularly in the East South Central division. Low–low clusters were concentrated in the upper Midwest and Mountain West.

### 3.2 Model Selection and Test Performance

Table 2 summarizes validation and held-out test performance for the four candidate models. Each partition contained 444 counties. Primary-model selection was based exclusively on validation-set RMSE. LightGBM achieved the lowest validation RMSE (0.4421), followed closely by XGBoost (0.4462), an absolute difference of 0.0041 percentage points. LightGBM was therefore designated as the primary model under the validation-based selection rule, and held-out test performance was reported without altering this decision. On the held-out test set, LightGBM achieved RMSE = 0.423 (95% CI: 0.374–0.482), MAE = 0.311 (95% CI: 0.287–0.338), MAPE = 2.76%, and *R*^2^ = 0.964. XGBoost achieved a modestly lower test RMSE of 0.399 (95% CI: 0.350–0.453) and *R*^2^ = 0.968 and was retained as a secondary benchmark. Random Forest and Elastic Net showed higher prediction errors. All pairwise Wilcoxon signed-rank tests comparing absolute test-set errors were statistically significant (all *p <* 0.01), including LightGBM versus XGBoost (*W* = 40,970, *p* = 0.0018). Observed-versus-predicted and residual plots for all four models are provided in S1 and S2 Figs.

### 3.3 Geographic Generalizability

Table 3 presents region-held-out validation results, in which each Census region was held out in turn while the model was trained on counties from the remaining three regions. This design provides a more conservative assessment of geographic generalizability than the stratified random split because it evaluates transfer to a geographically distinct region.

**Table 3:** Region-held-out validation results for the primary LightGBM model. Each Census region was held out in turn, and the model was trained on counties from the remaining three regions. Imputation and feature scaling were refitted within each fold using training counties only.

| Held-out region | $n$ counties | RMSE | MAE | MAPE (%) | $R^2$ |
| --- | --- | --- | --- | --- | --- |
| West | 450 | 0.733 | 0.584 | 6.13 | 0.831 |
| Midwest | 1,055 | 0.802 | 0.627 | 6.01 | 0.707 |
| South | 1,301 | 1.183 | 0.948 | 7.24 | 0.722 |
| Northeast | 151 | 0.995 | 0.882 | 10.11 | 0.336 |

**Table 4:** Sensitivity analysis results for the validation-selected LightGBM model. All specifications used the same selected LightGBM hyperparameters and were evaluated on the same held-out test set (*n* = 444 counties). The primary model is included for reference.

| Model | Specification | RMSE | MAE | MAPE (%) | $R^2$ |
| --- | --- | --- | --- | --- | --- |
| Primary | All 22 primary predictors | 0.423 | 0.311 | 2.76 | 0.964 |
| S1 | Structural and contextual predictors only | 0.923 | 0.673 | 5.97 | 0.827 |
| S2 | Health-behavior and clinical covariates only | 1.120 | 0.884 | 8.01 | 0.745 |
| S3 | FECI excluded; preprocessing refitted | 0.418 | 0.305 | 2.71 | 0.964 |
| S4 | Collinearity-reduced specification | 0.424 | 0.312 | 2.76 | 0.963 |
| S5 | Binary high-food-desert indicator | 0.424 | 0.308 | 2.74 | 0.963 |
| S6 | FECI excluded; primary preprocessing retained | 0.418 | 0.305 | 2.71 | 0.964 |
| S7 | Rurality excluded | 0.420 | 0.308 | 2.74 | 0.964 |
| S8 | Census-region indicators added | 0.407 | 0.291 | 2.57 | 0.966 |
| S9 | Top 10 training-ranked SHAP predictors | 0.400 | 0.296 | 2.63 | 0.967 |

Note: FECI, Food Environment Composite Index. Sensitivity specifications were used to assess robustness and were not treated as alternative primary-model candidates. Full specifications are provided in S4 Table.

Region-held-out validation used the primary LightGBM model with hyperparameters fixed after validation-based model selection. Within each regional fold, imputation and feature scaling were fitted using only counties from the three training regions and then applied to the held-out region. Performance was strongest when the West was held out (*R*^2^ = 0.831, RMSE = 0.733) and weakest when the Northeast was held out (*R*^2^ = 0.336, RMSE = 0.995). Performance was moderate for the South and Midwest, with all metrics reported in Table 3.

The weaker Northeast performance may partly reflect its smaller number of counties (*n* = 151), distinct predictor and outcome distributions, and limited representation of northeastern county profiles in the three training regions. Overall, the contrast between the stratified random test performance (*R*^2^ = 0.964) and the region-held-out results (*R*^2^ range: 0.336–0.831) indicates that random-split accuracy should not be interpreted as evidence of equivalent geographic transportability.

### 3.4 SHAP Feature Importance and Geographic Interpretation

Figure 2 presents the global SHAP feature importance ranking for all 22 primary predictors in the validation-selected LightGBM model. Physical inactivity prevalence had the highest mean absolute SHAP value (0.633 percentage points), followed by White population percentage (0.430), poverty rate (0.356), and Black population percentage (0.220). Among structural predictors, poverty rate, food insecurity rate, median household income, and SNAP participation rate were the highest-ranked contributors. The food desert index and Food Environment Composite Index had lower global SHAP importance, indicating limited incremental contribution after accounting for broader socioeconomic, demographic, behavioral, and clinical predictors.

**Figure 1:**
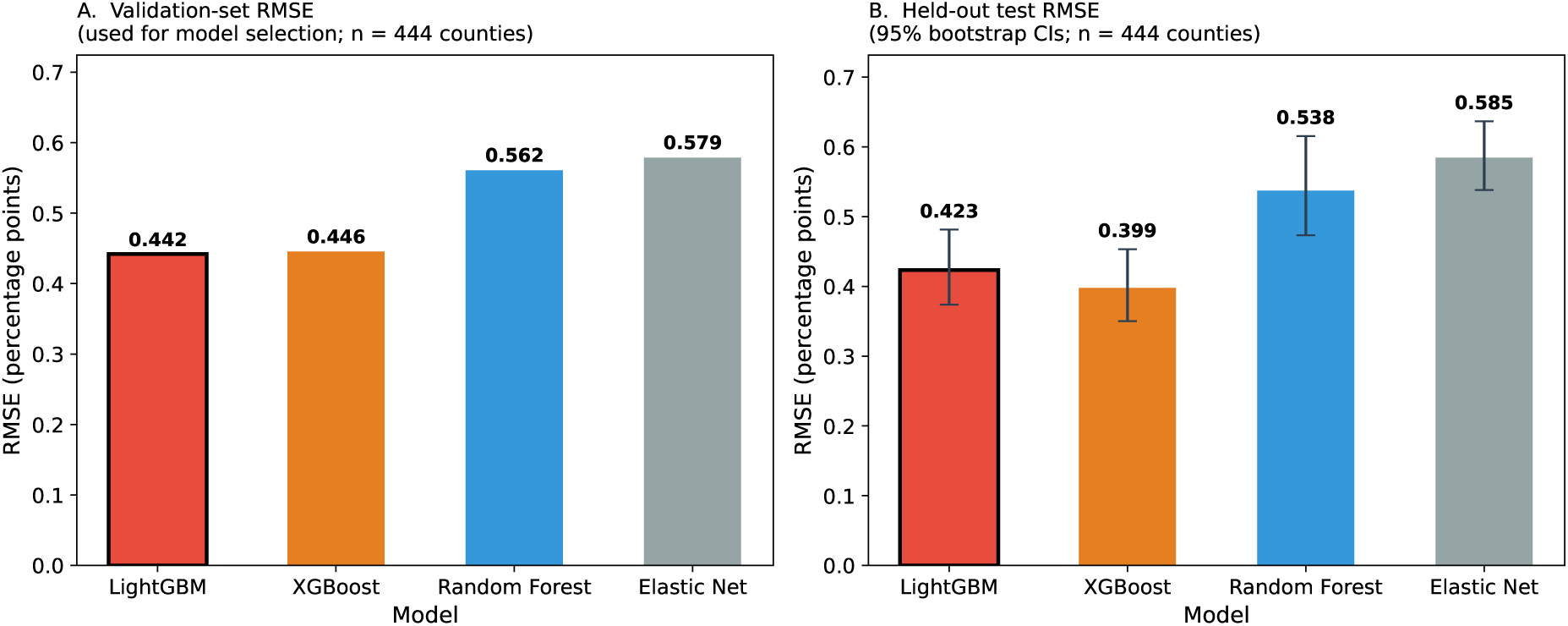
Validation-based model selection and held-out test performance. (A) Validation-set RMSE for the four candidate models. LightGBM achieved the lowest validation RMSE and was selected as the primary model. (B) Held-out test-set RMSE with 95% bootstrap confidence intervals. Although XGBoost achieved the lowest test RMSE, LightGBM remained the primary model because model selection was based exclusively on validation performance. The black outline identifies LightGBM as the validation-selected primary model.

**Figure 2:**
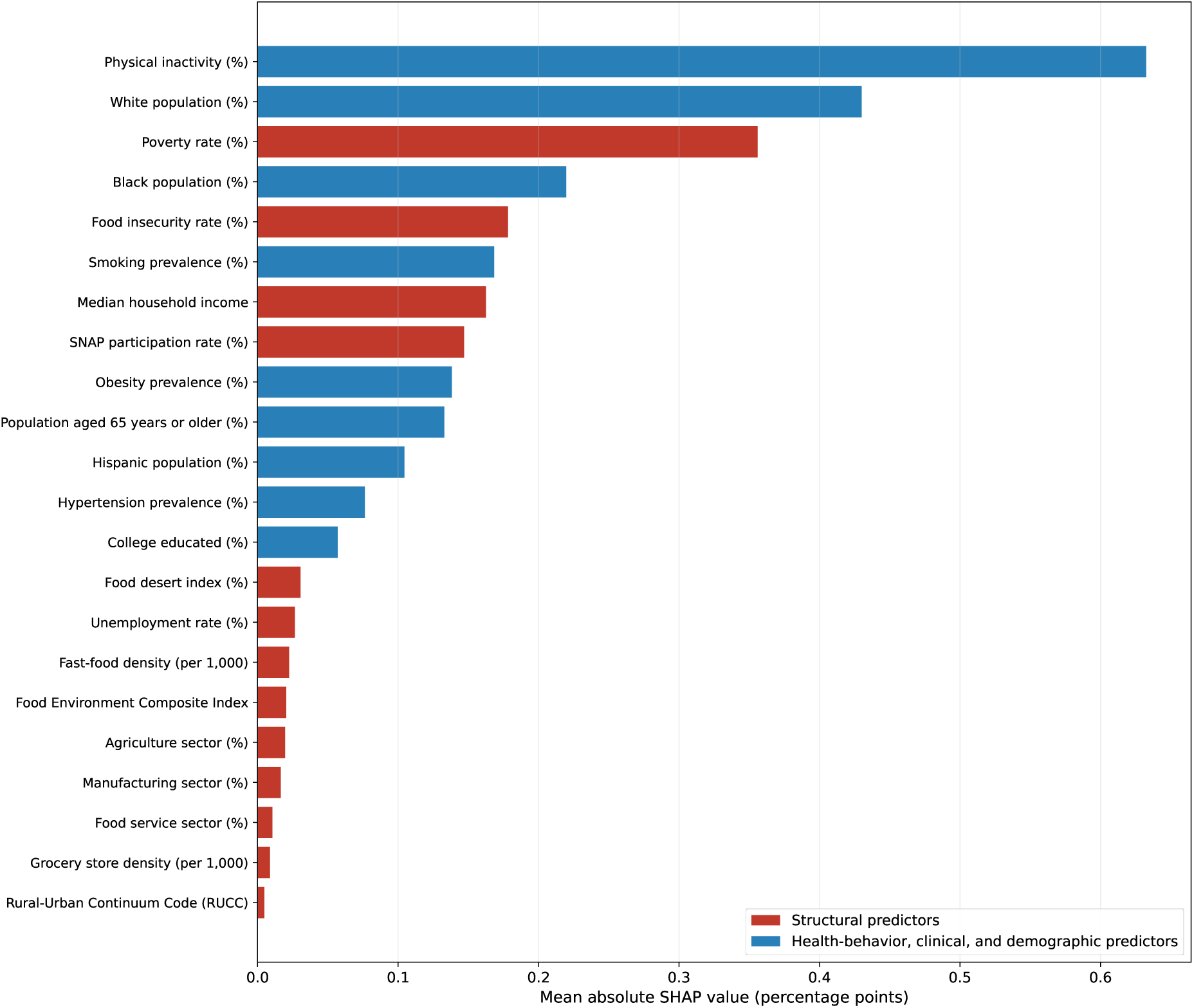
Global feature importance in the validation-selected LightGBM model (*n* = 2,957 counties). Bars show all 22 primary predictors ranked by mean absolute SHAP value. Larger values indicate greater average contribution magnitude but do not indicate direction or causal effects. Red bars represent structural predictors, and blue bars represent health-behavior, clinical, and demographic predictors. SHAP values are expressed in percentage points of predicted county-level diagnosed diabetes prevalence.

County-level racial and ethnic composition variables were interpreted as contextual markers of racialized social, economic, and geographic inequities rather than as biological risk factors. Mean absolute SHAP values summarize average contribution magnitude and do not indicate direction or causality. Directional feature contributions are shown in the SHAP summary plot (Figure 3).

**Figure 3:**
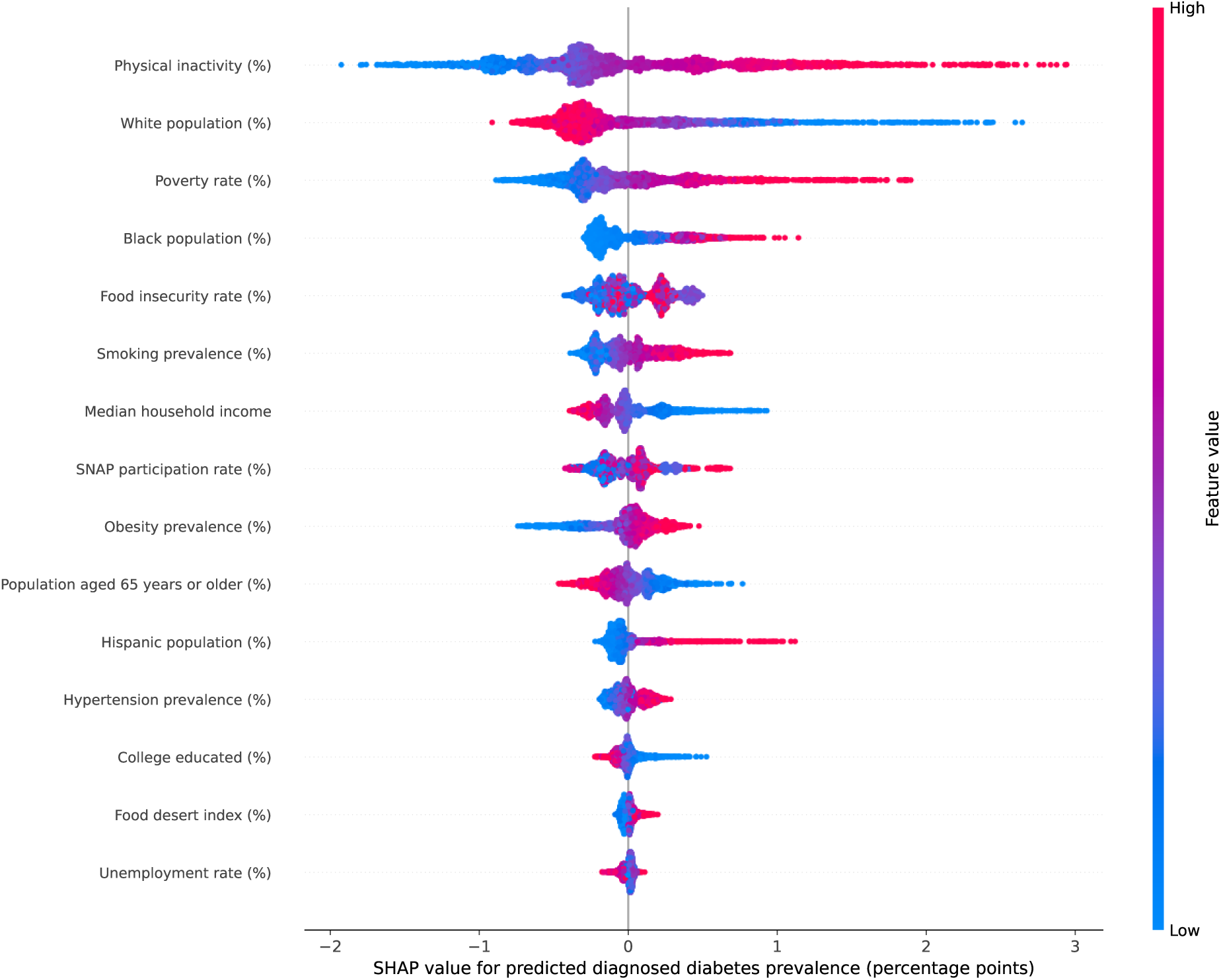
SHAP summary plot for the validation-selected LightGBM model. The figure shows the 15 predictors with the highest mean absolute SHAP values across all 2,957 counties. Each point represents one county, and color indicates the relative scaled feature value. Positive SHAP values indicate contributions toward higher predicted diagnosed diabetes prevalence, whereas negative values indicate contributions toward lower predicted prevalence. SHAP values explain model predictions only and should not be interpreted as causal effects.

A cross-model agreement analysis showed that LightGBM and XGBoost produced highly consistent global explanations (Spearman’s *ρ* = 0.988, *p <* 0.001), shared four of the five highest-ranked predictors, and generated strongly correlated county-level predictions (Pearson’s *r* = 0.998). The models assigned the same dominant positive structural SHAP contributor to 76.5% of counties, indicating substantial but incomplete agreement in local geographic explanations.

County-level SHAP maps for poverty rate, SNAP participation rate, and median household income showed distinct geographic patterns (S3–S5 Figs), and corresponding SHAP dependence plots are provided in S6–S8 Figs. Poverty rate made strong positive contributions in many counties across the Deep South and Appalachia, SNAP participation showed more geographically heterogeneous contributions, and median household income generally contributed positively at lower income levels and negatively at higher income levels.

For the dominant positive structural contributor analysis, the structural predictor with the largest positive SHAP contribution was identified for each county. Poverty rate was the most common dominant positive structural contributor (*n* = 772 counties, 26.1%), followed by food insecurity rate (*n* = 707, 23.9%), SNAP participation rate (*n* = 316, 10.7%), unemployment rate (*n* = 224, 7.6%), median household income (*n* = 178, 6.0%), and food desert index (*n* = 162, 5.5%). Other structural predictors accounted for 597 counties (20.2%), and one county had no positive structural SHAP contribution.

Dominant positive structural contributors varied by region. Poverty rate and food insecurity rate were most prominent in the South, food insecurity rate and poverty rate were most common in the Midwest, poverty rate and SNAP participation rate were most common in the West, and unemployment rate was most common in the Northeast. The complete within-region distribution is shown in S10 Fig. These dominant contributors describe model predictions rather than causal effects or expected intervention benefits and should be interpreted alongside local surveillance data, stakeholder knowledge, and causal evidence. Regional mean signed SHAP profiles are provided in S11 Fig.

### 3.5 Spatial Autocorrelation Diagnostics

Observed county-level diagnosed diabetes prevalence showed strong positive spatial autocorrelation (Global Moran’s *I* = 0.665, permutation *p* = 0.001, queen contiguity). After fitting the validation-selected LightGBM model, residual spatial autocorrelation decreased to *I* = 0.069 (*p* = 0.001), corresponding to an approximately 89.6% relative reduction in Moran’s *I*. Although the remaining residual autocorrelation was small in magnitude, it remained statistically significant, indicating that some geographically structured variation was not captured by the included predictors. The complete county-level residual choropleth is provided in S9 Fig.

The validation-selected LightGBM model fitted on the training partition was applied to all 2,957 analytic counties to generate predictions and residuals. Residual Moran’s *I* was calculated using the 2,912 counties whose identifiers matched the shapefile geometries. Residuals for training counties were therefore in-sample, whereas residuals for validation and test counties were out-of-sample.

Local Indicators of Spatial Association identified 470 high–high clusters, 647 low–low clusters, 28 low–high spatial outliers, and 30 high–low spatial outliers among the 2,912 spatially matched counties. The remaining 1,737 counties had no statistically significant local spatial association at the unadjusted permutation threshold of *p <* 0.05. High–high clusters were concentrated primarily in the South, whereas low–low clusters were concentrated in the upper Midwest and parts of the Mountain West (Figure 5). These LISA results were interpreted as exploratory because local tests were not adjusted for multiple comparisons.

**Figure 4:**
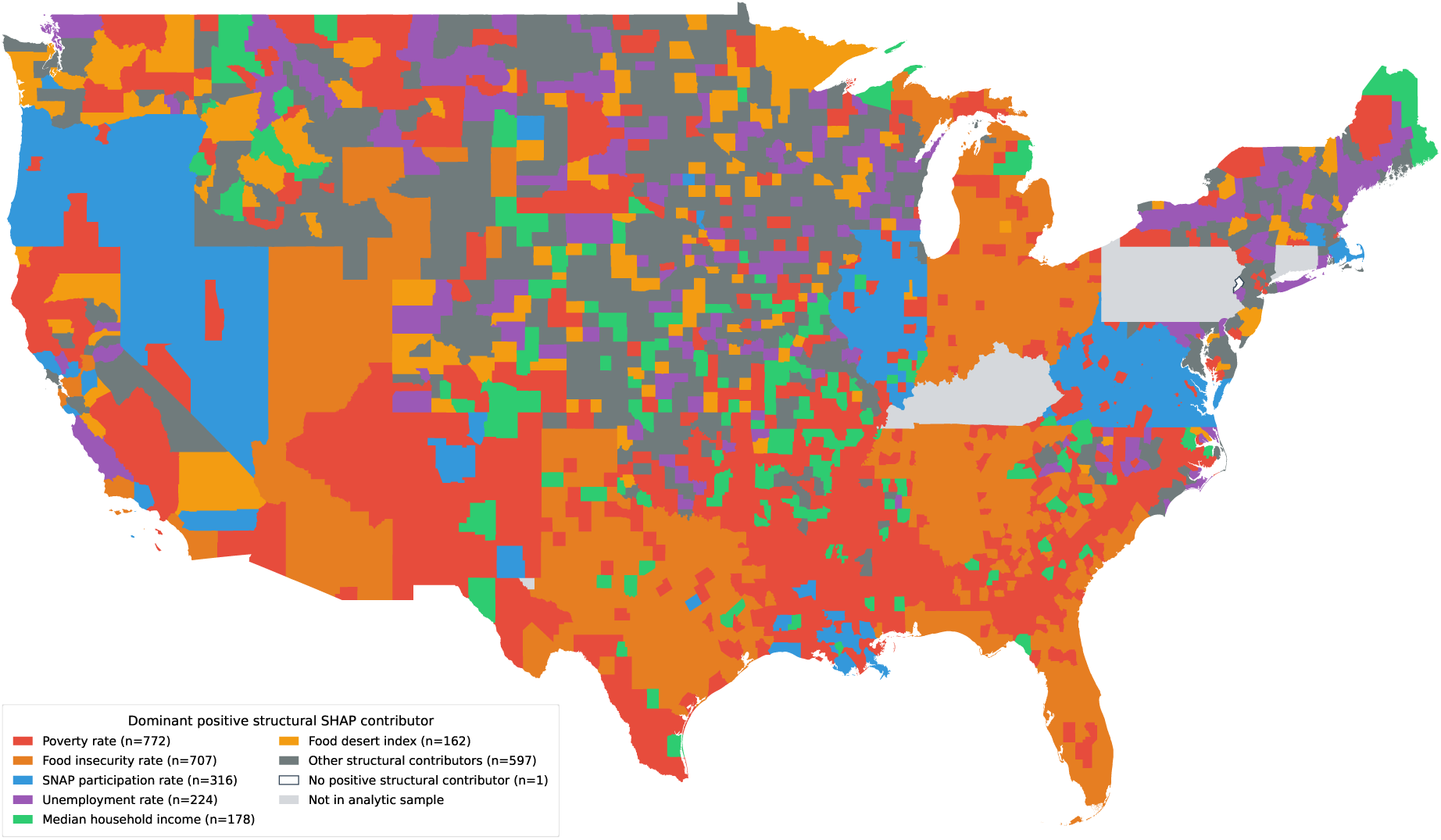
Dominant positive structural SHAP contributor by county in the validation-selected LightGBM model. For each county with at least one positive structural SHAP value, the structural predictor with the largest positive contribution was identified. Frequencies shown in the legend are based on all 2,957 analytic counties; the map displays the contiguous United States. Counties shown in light gray were not included in the analytic sample. Dominant contributors explain model predictions and should not be interpreted as causal determinants or expected intervention effects.

**Figure 5:**
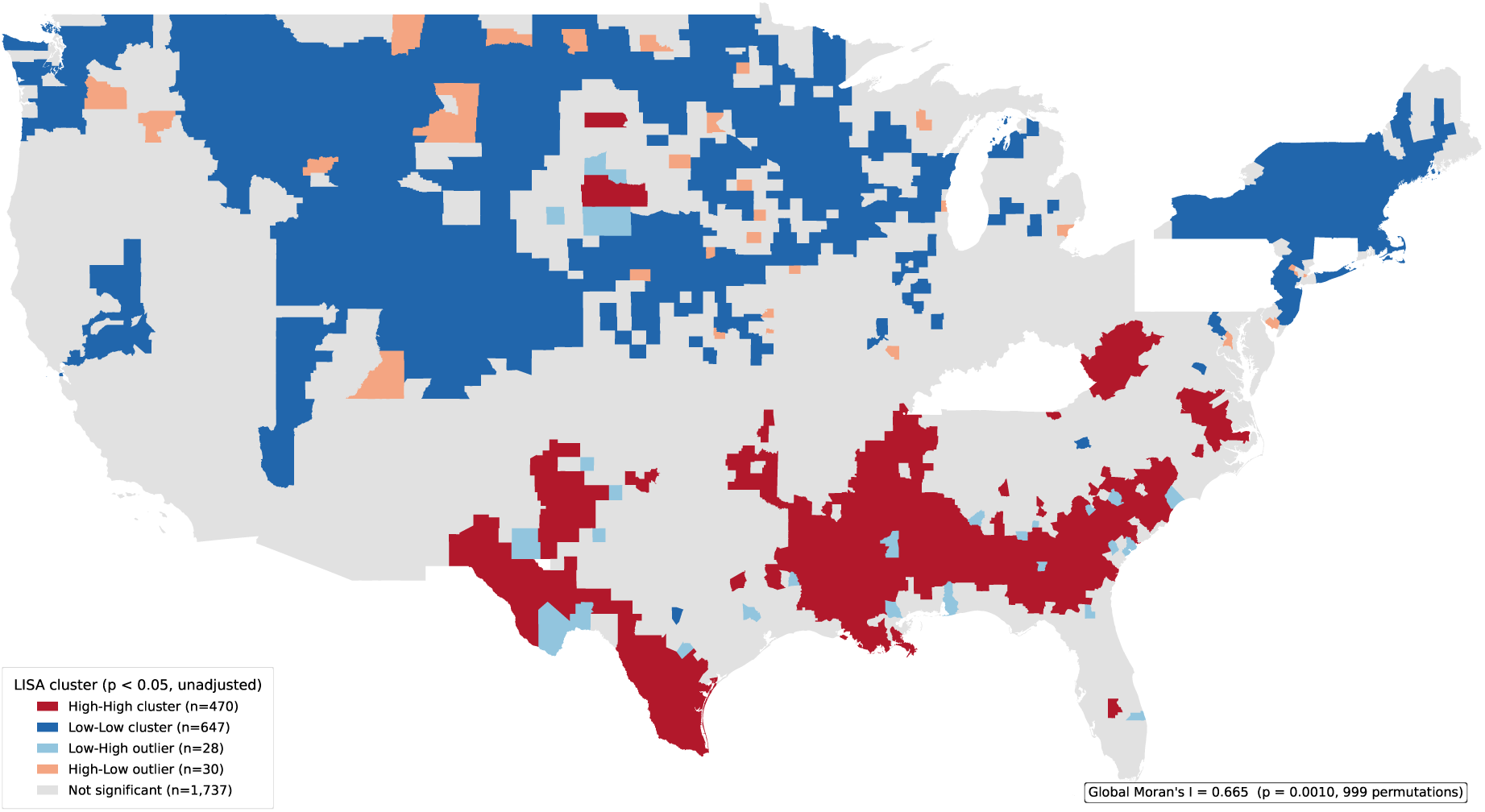
Local spatial clusters of observed county-level diagnosed diabetes prevalence among the 2,912 spatially matched counties. Local Moran’s *I* was calculated using row-standardized queen-contiguity weights and 999 permutations. High–high clusters indicate counties with high prevalence surrounded by counties with high prevalence, whereas low–low clusters indicate counties with low prevalence surrounded by counties with low prevalence. Low–high and high–low categories represent spatial outliers. Local statistical significance was evaluated using an unadjusted threshold of *p <* 0.05; results are exploratory.

### 3.6 Sensitivity Analyses

Table 4 presents the results of nine sensitivity specifications. All sensitivity models used the validation-selected LightGBM architecture with the same selected hyperparameters as the primary model and were evaluated on the same held-out test set (*n* = 444 counties). These analyses were treated as robustness checks and were not considered alternative candidates for primary-model selection.

The structural-and-contextual-predictors-only model (S1) retained substantial predictive performance (*R*^2^ = 0.827), whereas the health-behavior-and-clinical-covariates-only model (S2) performed less well (*R*^2^ = 0.745). This indicates that structural and contextual county-level predictors contained considerable predictive information even without CDC PLACES health-behavior and clinical covariates, while the full model benefited from combining both predictor groups.

Model performance was robust to several alternative specifications. Excluding the Food Environment Composite Index, with preprocessing refitted (S3) or retained from the primary pipeline (S6), produced the same rounded performance (*R*^2^ = 0.964). The collinearity-reduced model (S4), binary high-food-desert model (S5), and rurality-excluded model (S7) also performed similarly to the primary model. Adding Census-region indicators (S8) and using the 10 highest-ranked training-partition SHAP predictors (S9) yielded modestly lower test errors, but these were post-selection robustness analyses and did not revise the validation-based primary-model selection. Overall, predictive performance was not materially dependent on FECI, rurality, the continuous food-desert formulation, the identified collinear predictors, or the full 22-predictor feature set.

## 4 Discussion

### 4.1 Principal Findings

This study developed an explainable gradient-boosting framework for predicting county-level diagnosed diabetes prevalence across 2,957 U.S. counties using publicly available structural, food-environment, socioeconomic, demographic, health-behavior, and clinical indicators. Three principal findings emerged. First, the validation-selected LightGBM model achieved strong performance under the stratified random county split (*R*^2^ = 0.964, RMSE = 0.423 percentage points), while XGBoost achieved slightly lower test error but was retained as a secondary comparator because primary-model selection was based on validation-set RMSE. Region-held-out validation showed weaker transportability, especially in the Northeast (*R*^2^ = 0.336), indicating that random-split performance should not be interpreted as equivalent geographic generalizability.

Second, structural and contextual predictors retained substantial predictive information even without the CDC PLACES health-behavior and clinical covariates. The structural-and-contextual-predictors-only model achieved *R*^2^ = 0.827, whereas the health-behavior-and-clinical covariates-only model achieved *R*^2^ = 0.745. The full model outperformed both restricted specifications, suggesting that structural/contextual and health-related predictor groups provided complementary information for predicting county-level diagnosed diabetes prevalence. These findings are consistent with prior research linking food insecurity, economic disadvantage, and neighborhood conditions with diabetes burden [11, 18].

Third, SHAP-based geographic interpretation showed that the structural predictors making the largest positive contributions to model predictions varied across counties and Census regions. Poverty rate was the most frequent dominant positive structural SHAP contributor nationally, followed by food insecurity rate, SNAP participation rate, unemployment rate, median household income, and food desert index. Within-region patterns were heterogeneous: poverty rate was most common in the South and West, food insecurity rate was most common in the Midwest, and unemployment rate was most common in the Northeast. These dominant contributors identify which structural predictors most increased the fitted model’s prediction for each county; they do not identify causal determinants or the interventions expected to produce the largest reductions in diabetes prevalence.

### 4.2 Structural Predictors and Model Robustness

The global SHAP ranking indicated that broader socioeconomic and food-security measures contributed more strongly to model predictions than several narrower food-access and retail-density indicators. Poverty rate, food insecurity rate, median household income, and SNAP participation rate ranked among the highest structural contributors, whereas the food desert index, fast-food density, Food Environment Composite Index, grocery store density, and rurality had lower global SHAP importance. This does not imply that food access or rurality are unimportant to diabetes burden. Rather, within this national county-level prediction model, these measures contributed less incremental predictive information after accounting for poverty, income, food insecurity, demographic context, health-behavior indicators, and other county-level variables.

The sensitivity analyses supported the robustness of this interpretation. Excluding the Food Environment Composite Index before preprocessing refitting (S3) or after primary preprocessing (S6) produced the same rounded performance (*R*^2^ = 0.964), indicating that the composite added little incremental predictive information beyond its component variables and the broader predictor set. The collinearity-reduced model, rurality-excluded model, and binary high-food-desert specification also performed similarly to the primary model. Census-region indicators and the reduced top-10 predictor model produced slightly lower test errors, but these were post-selection robustness checks and did not revise the validation-based primary-model selection.

The comparison of LightGBM and XGBoost further suggested that the main findings were not unique to one gradient-boosting implementation. The two models produced highly consistent global SHAP rankings (Spearman’s *ρ* = 0.988), strongly correlated county-level predictions (Pearson’s *r* = 0.998), and the same dominant positive structural SHAP contributor in 76.5% of counties. This agreement supports the stability of the global findings, while the incomplete county-level agreement indicates that some local explanations remained model dependent.

### 4.3 Geographic Interpretation, Causality, and Equity

The SHAP analysis extends prior explainable-AI work in diabetes research [12] from individual-level clinical interpretation to county-level geographic prediction. County-level SHAP maps and dominant-contributor summaries may help identify places and contextual conditions requiring further investigation. For example, counties where poverty or food insecurity made large positive contributions may warrant closer examination of economic conditions, food-security programs, healthcare access, and local service availability. Counties where unemployment was prominent may warrant investigation of income instability and its relationship with food access and healthcare utilization. These applications should be viewed as hypothesis-generating and should be combined with local surveillance data, stakeholder knowledge, program-effectiveness evidence, and causal research.

The geographic explanations require careful interpretation. SHAP values describe how predictors contributed to fitted model predictions relative to the model baseline; they do not show that changing a predictor would produce a corresponding change in diagnosed diabetes prevalence [24]. The racial and ethnic composition variables require particular caution. Their SHAP contributions should not be interpreted as biological effects or intrinsic population risks. Instead, county-level racial and ethnic composition may reflect historically structured differences in residential segregation, economic opportunity, environmental exposure, healthcare access, food environments, and other social conditions. The ecological design also precludes individual-level inference; county associations cannot be assumed to represent individual exposure–risk relationships.

### 4.4 Limitations

Several limitations should be considered. First, the ecological cross-sectional design precludes causal inference and does not establish temporal ordering among predictors and diagnosed diabetes prevalence. Diagnosed diabetes prevalence in CDC PLACES is based on self-reported BRFSS data and multilevel regression and poststratification, and may underestimate the true burden in areas where undiagnosed diabetes is common or healthcare access is limited [1]. In addition, the outcome and the four CDC PLACES health-behavior and clinical covariates were derived from the same BRFSS-based surveillance and estimation system, which may have increased apparent predictive performance. The structural-and-contextual-predictors-only sensitivity model reduced this concern by showing substantial performance without those same-source covariates, but it did not eliminate all potential dependence among variables.

Second, several predictors were imperfect proxies for complex local conditions. The USDA food-desert measure captures geographic proximity to supermarkets but does not account for food quality, affordability, transportation access, online grocery delivery, mobile food markets, or household-level purchasing behavior [10]. Food retail density also does not directly measure whether residents can afford or use available food resources. Similarly, occupational-sector shares from the ACS describe the formally employed population and may not adequately represent informal, temporary, or gig-economy workers [18]. The analytic sample excluded county-equivalent units that could not be consistently linked across all data sources, and Pennsylvania and Kentucky counties were absent from the CDC PLACES county-level file used in this analysis. Spatial analyses were further limited to 2,912 of the 2,957 analytic counties because 45 county identifiers could not be matched to the geometry file. LISA clusters were identified using an unadjusted permutation threshold of *p <* 0.05 and should therefore be interpreted as exploratory.

Third, some model-based findings require cautious interpretation. Residual spatial autocorrelation remained statistically significant after LightGBM fitting (Global Moran’s *I* = 0.069, *p* = 0.001), indicating that some geographically structured variation was not captured by the included predictors. Region-held-out validation showed weaker geographic transportability, especially in the Northeast (*R*^2^ = 0.336, RMSE = 0.995), and temporal stability was not tested against future CDC PLACES releases. The Food Environment Composite Index was constructed using PCA fitted on the full analytic sample before data partitioning, creating limited unsupervised preprocessing leakage; although FECI-exclusion analyses suggested little influence on primary test performance, refitting PCA within each training partition would provide a more rigorous assessment. Finally, SHAP values can distribute attribution among correlated predictors, and county-level dominant-contributor assignments may vary across model specifications.

Despite these limitations, the study demonstrates that explainable gradient boosting, public county-level data, and spatial diagnostics can be combined to characterize geographic variation in diagnosed diabetes prevalence. The framework offers a reproducible approach for identifying predictive patterns and generating local public-health hypotheses while maintaining a clear distinction between model explanation and causal inference.

## 5 Conclusion

This study developed an explainable gradient-boosting framework for predicting county-level diagnosed diabetes prevalence in the United States using publicly available food-environment, socioeconomic, occupational, demographic, health-behavior, and clinical indicators. LightGBM was selected as the primary model based on validation-set RMSE and achieved strong held-out test performance (*R*^2^ = 0.964; RMSE = 0.423 percentage points). However, weaker region-held-out performance indicated that random-split accuracy should not be interpreted as equivalent geographic transportability.

The structural-and-contextual-predictors-only sensitivity model retained substantial predictive performance (*R*^2^ = 0.827) after excluding CDC PLACES health-behavior and clinical covariates, indicating that county-level socioeconomic, demographic, food-environment, occupational, and rurality indicators contained considerable predictive information. SHAP-based analyses translated model predictions into county-level and regional explanations. Poverty rate and food insecurity rate were the most frequent dominant positive structural contributors nationally, while SNAP participation, unemployment, median household income, food desert index, and other structural variables were prominent in particular geographic contexts.

These geographic explanations may support local hypothesis generation and help identify contextual conditions requiring further investigation. They should not be interpreted as causal determinants, intervention effects, or evidence that the dominant predictor is necessarily the best target for resource allocation. Application of the maps should be combined with local surveillance data, stakeholder knowledge, program-effectiveness evidence, and causal research.

Future work should evaluate temporal stability using longitudinal county panels, future CDC PLACES releases, and independent state- or county-level surveillance systems. Spatial block validation, state-held-out validation, region-specific calibration, subgroup performance analyses, and more detailed measures of food affordability, transportation, healthcare access, digital food access, and local policy context would further strengthen geographic and equity-related model assessment. Overall, the framework is best viewed as a predictive and hypothesis-generating tool rather than as a causal decision system.

## Ethics Statement

This study used only publicly available, aggregate county-level data from federal public data sources. No individual-level records, personally identifiable information, or private human-subjects data were accessed. Institutional review board approval was not required.

## Data Availability

All raw input data used in this analysis are publicly available from CDC PLACES, the USDA Economic Research Service, the U.S. Census Bureau American Community Survey, and the USDA Rural–Urban Continuum Codes. The processed county-level analytic dataset, Python analysis code, model outputs, and figure-generation scripts needed to reproduce the findings are archived in Zenodo: https://doi.org/10.5281/zenodo.20801697. The corresponding GitHub repository is available at: https://github.com/YUSSIF47/county-diabetes-lightgbm-shap.

## Funding Statement

The authors received no specific funding for this work.

## Competing Interests

The authors declare no competing interests.

## Author Contributions

Conceptualization: Y.Y., S.K., P.R.S., M.A.A.M.

Data curation: P.R.S., M.A.A.M.

Formal analysis: Y.Y., S.K.

Methodology: Y.Y., P.R.S., S.K.

Software: Y.Y.

Visualization: Y.Y.

Writing – original draft: P.R.S., Y.Y.

Writing – review & editing: P.R.S., S.K., Y.Y., M.A.A.M.

## Acknowledgments

The authors thank the Centers for Disease Control and Prevention, the USDA Economic Research Service, and the U.S. Census Bureau for making the data sources used in this study publicly available. Computational analyses were performed on the UTRGV CRA-DLE high-performance computing cluster.

## Supporting Information

The supporting information file contains supplementary tables and figures, including missingness summaries, variance inflation factor diagnostics, hyperparameter search grids, sensitivity-analysis specifications, observed-versus-predicted plots, residual plots, county-level SHAP maps, SHAP dependence plots, the residual choropleth map, the within-region distribution of dominant positive structural SHAP contributors, and regional mean signed SHAP profiles.

## Notes

### Competing Interest Statement

The authors have declared no competing interest.

### Funding Statement

The author(s) received no specific funding for this work.

### Author Declarations

This study used only publicly available, aggregate county-level data from public data sources. The analysis did not involve individual-level records, personally identifiable information, private health information, clinical intervention, recruitment, or direct contact with human participants. Therefore, formal IRB approval was not required.

